# A retrospective study of the effects of COVID-19 Non-pharmaceutical interventions on Influenza in Canada

**DOI:** 10.1101/2024.02.16.24302930

**Authors:** Matthew I Betti, Heather MacTavish, Kenzie MacIntyre, Paniz Zadeh

## Abstract

The COVID-19 pandemic had a significant impact on endemic respiratory illnesses. Through behavioural changes in populations and government policy, mainly through non-pharmaceutical interventions (NPIs), Canada saw historic lows in the number of Influenza A cases from 2020 through 2022. In this study, we use historical influenza A data for Canada and three provincial jurisdictions within Canada: Ontario, Quebec and Alberta to quantify the effects of these NPIs on influenza A. We aim to see which base parameters and derived parameters of an SIR model are most affected by NPIs. We find that the effective population size is the main driver of change, and discuss how these retrospective estimates can be used for future forecasting.

## 1. Introduction

The COVID-19 pandemic had a substantial impact on every facet of daily life around the world. In Canada, the non-pharmaceutical interventions (NPIs) put in place for much of the time period beginning around March 2020 to approximately the end of 2022 had a profound impact on other endemic respiratory infections.

Beginning in March 2020, jurisdictions across Canada experienced a variety of measures that could impact the spread of airborne infectious diseases. These measures included lockdowns (1, 2), masking (both on a personal and institutional level (3, 4), and social distancing (5). While adherence, enforcement and austerity of these measures in Canada differed by province (6), they left a measurable effect on seasonal Influenza.

Almost immediately, as NPIs took hold a measurable effect was seen on Influenza testing as reported by FluWatch (7). In fact, it is possible that one of the two major lineages of Influenza B may have been driven to extinction through COVID-19 and associated NPIs (8). In this study, we hope to measure the qualitative and quantitative effects of NPIs on Influenza A and B in Canada.

Influenza has long and thoroughly been studied using mathematical modeling. Using compartmental models, to study the spread of infectious diseases has been used for over a century, with work being done as early as 1911 (9). The compartmental SIR framework, where a population is broken into three classes: (S)usceptible, (I)nfectious, and (R)ecovered, is a popular choice for influenza modeling (10) and can be easily adapted to study different aspects of an epidemic such as cross-immunity from previous infections(11), vaccination strategies (12),. The SIR model is often well suited for parameter estimation of key epidemiological parameters, given appropriate population level data (13).

Unfortunately, testing and reporting is often biased toward severe cases (14), and therefore will severely underestimate the true number of cases within a population. Since 2020, the amount of data collected - particularly for SARS-CoV-2 infections - has improved leading to better estimates of parameters (15). Meanwhile, at least in Canada, the collection and reporting of influenza has not changed. While this can provide overestimates or underestimates of model parameters (16), in this study we hope to circumvent the limits of available data by focusing on relative comparisons of parameters.

Here we use a simple SIR model coupled with influenza A data from Summer 1999 to Spring 2023 to estimate relative changes in the reproduction number, attack rate, effective susceptible population, and contact rate induced by NPIs and COVID-19 policy. An SIR model is chosen for its simplicity as influenza data is reported weekly, resulting in fewer than 50 data points per season. We discuss the interpretation and impacts of these parameter estimates and changes in the context of other endemic or emerging infectious diseases, and in relation to policy.

## 2. Methodology

In this study, in order to reduce the effects of correlations, we will use a simple SIR model with our historical flu data. This technique is not new, and is the basis for forecasting of seasonal influenza in some studies (16, 17) and is used to simulate data in other cases (18). In implementing this model, we reduce the number of parameters required to fit by fixing those that are less likely to change over the period of time we are considering (i.e. the rate of recovery). The effective starting susceptible population is highly variables year-to-year as it depends on vaccination rates, vaccine efficacy, social behaviours, weather, etc. We fit this parameter and show that NPIs largely affect the effective population size.

Our model framework is

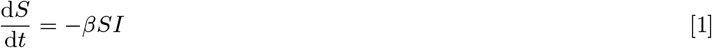

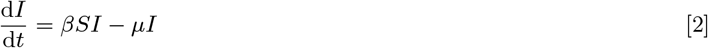

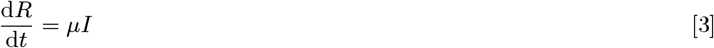

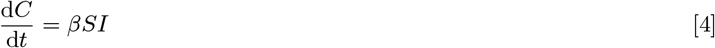

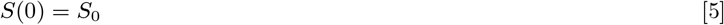

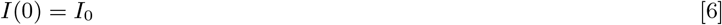

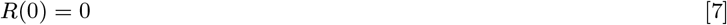

where *S* is the susceptible population, *I* is the infected population and *R* are those that have recovered with immunity. The model is augmented with *C*, the cumulative case counts for the flu season. We do this so that we can fit both *dC/dt* (new cases per week) and *C*(*t*) (cumulative cases) to data.

While *S*_0_ is often taken to be the whole population, this is complicated for influenza due to potential cross-immunity (11) and vaccination(19). We take *S*_0_ then to be the effective susceptible population size at the beginning of the flu season. In the context of our study, this is further complicated as NPIs effectively remove individuals from the population in some capacity. Thus, we consider *S*_0_ to be an *effective* initial susceptible population.

We likewise assume that the demographic changes within the approximate six months of the influenza season are negligible. These assumptions are again made to reduce the number of parameters of the model (to reduce covariance) and to keep the model mathematically tractable.

We estimate a seasonal reproduction number for each flu season using the standard expression for the reproduction number of an *SIR* model.

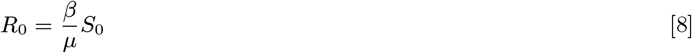

We do not start each year with a fully susceptible population, and thus this is not a true basic reproduction number (20). Our interpretation of *S*_0_ as an effective population size, leads to the interpretation of *R*_0_ as a seasonal reproduction number. In other words, it measures the effective reproduction of Influenza A at the start of a season.

We use provincial and national influenza data from Canada obtained from the publicly available FluWatch (21). Our interest is mainly in how model parameters differ in the 2020-2021 and 2021-2022 flu seasons.

## 3. Fitting

We fit this model using least squares to minimize the error between the cumulative case counts per week and the new cases per week. We use both data sets as often corrections and delays in data collection and reporting will be reflected in cumulative case counts but not in the weekly new case counts. By using both, we are able to minimize the effects of errors in data collection/reporting.

By using both cumulative cases and new reported cases, we also are able to take into consideration the different scales of reported cases across an outbreak. If we only used cumulative case data for instance, our fitting would favour the later points in an outbreak as the magnitude of these points will create a larger difference in the residual. We show in Figure 1 that this method does indeed produce a small relative error as well 9, indicating good fits to the beginning and end of an outbreak.

**Fig. 1.**
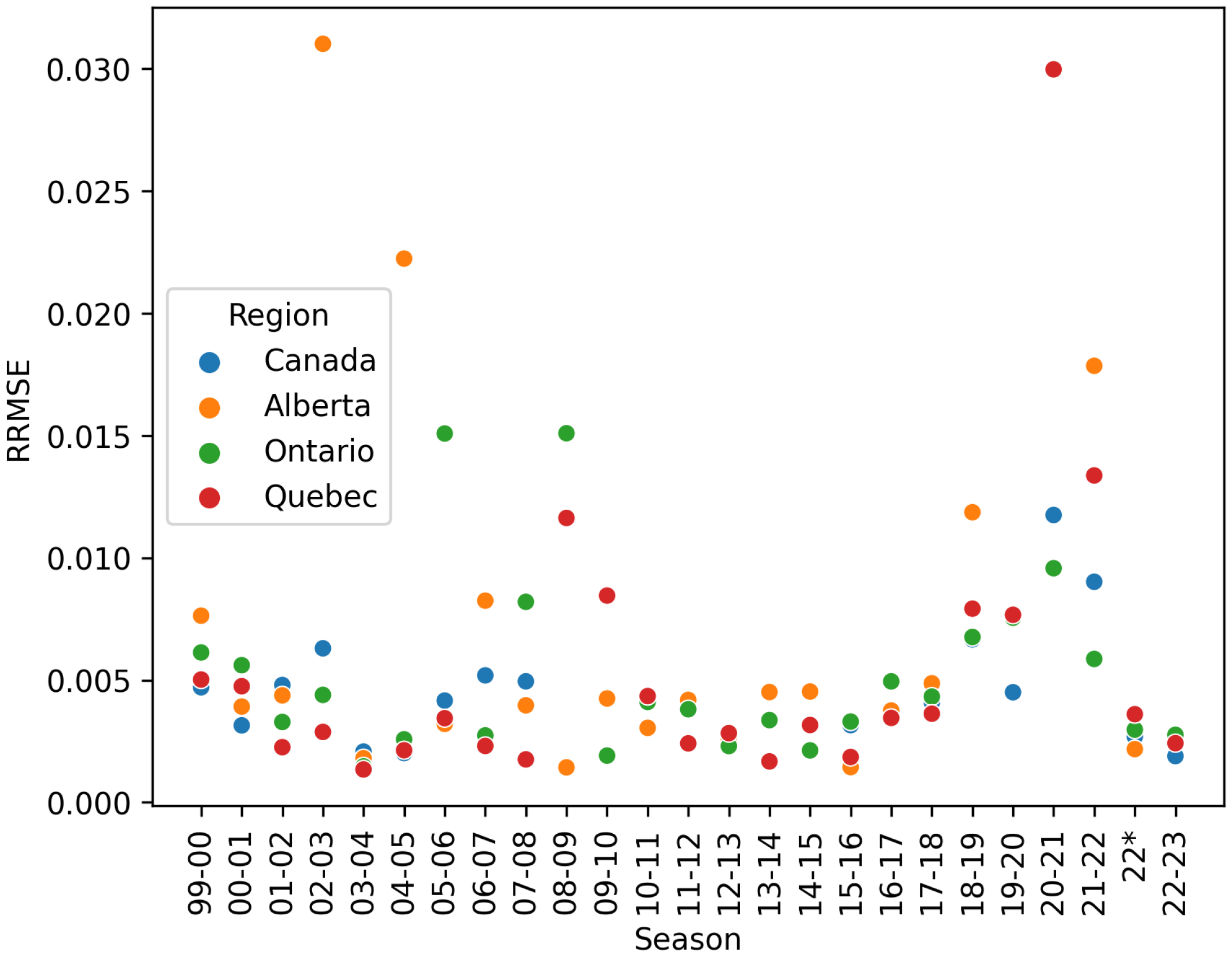
Root Mean-Square Error, equation Eq. (9) for the best fit model for each season. We use the RRMSE and a visual inspection to flag and remove bad or non-convergent fits.

We fix *μ* = 7*/*2 based on established data that the flu lasts approximately 2.5 days (19, 22, 23). We do this as we do not expect non-pharmaceutical interventions to change the rate of recovery of individuals within the population.

An influenza season is defined approximately from August of a given calendar year until July of the following calendar year. As an example, the 99-00 season runs from August 1999 through July, 2000.

Seasons are fit sequentially against data from Canada’s FluWatch (21) where available. For Ontario and Quebec, provincial level databases, (24) and (25), respectively, were used when FluWatch data was missing. We use direct reported case values for cumulative cases and new cases as opposed to percent positivity.

Literature estimates are used for the initial guess at *β* (19).

The initial susceptible population, *S*_0_, is fit. We use the total number of tests done in a season as a starting point for fitting; where this is unavailable, we use an estimated attack rate of 0.25 (19) and start fitting from the cumulative cases at the end of the season over the assumed attack rate.

Starting values and fixed values for parameter fitting are given in Table 1.

**Table 1.** Table of initial values for model fitting.

| Parameter | Value | Fixed |
| --- | --- | --- |
| $\beta$ | 0.000017 | False |
| $S_0$ | Total Seasonal Tests or Estimated | False |
| $\mu$ | 7/2.5 | True |
| $I(0)$ | 1 | False |
| $R(0)$ | 0 | True |

This method of fitting give the lowest relative root-mean square error,

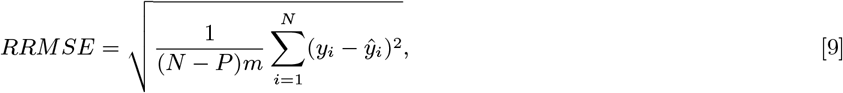

where *N* is the number of of data points, *P* is the number of parameters fit, *m* is the mean of squares of all values fit, *y*_*i*_ are the data points and *ŷ*_*i*_ are the fitted values.

We compare two fitting algorithms per season: least-squares and dual annealing, and take the better fit of the two in a relative root-mean-square error sense.

## 4. Results

Figure 1 shows the relative root mean-square error of the fit for each season. We can see from the plot that we generally fit seasonal data quite well. Any fit with a RMSE *>* 2% is discounted as the fitting did not visually accurately describe the data in any of these cases.

Figure 2 shows these fits overlaid on the data for each season for Canada. Due to each season being considered in isolation, there is little continuity between the seasons. We see that the fitted curves generally agree with the data on a per-season basis. Provincial level fits give qualitatively similar visuals.

**Fig. 2.**
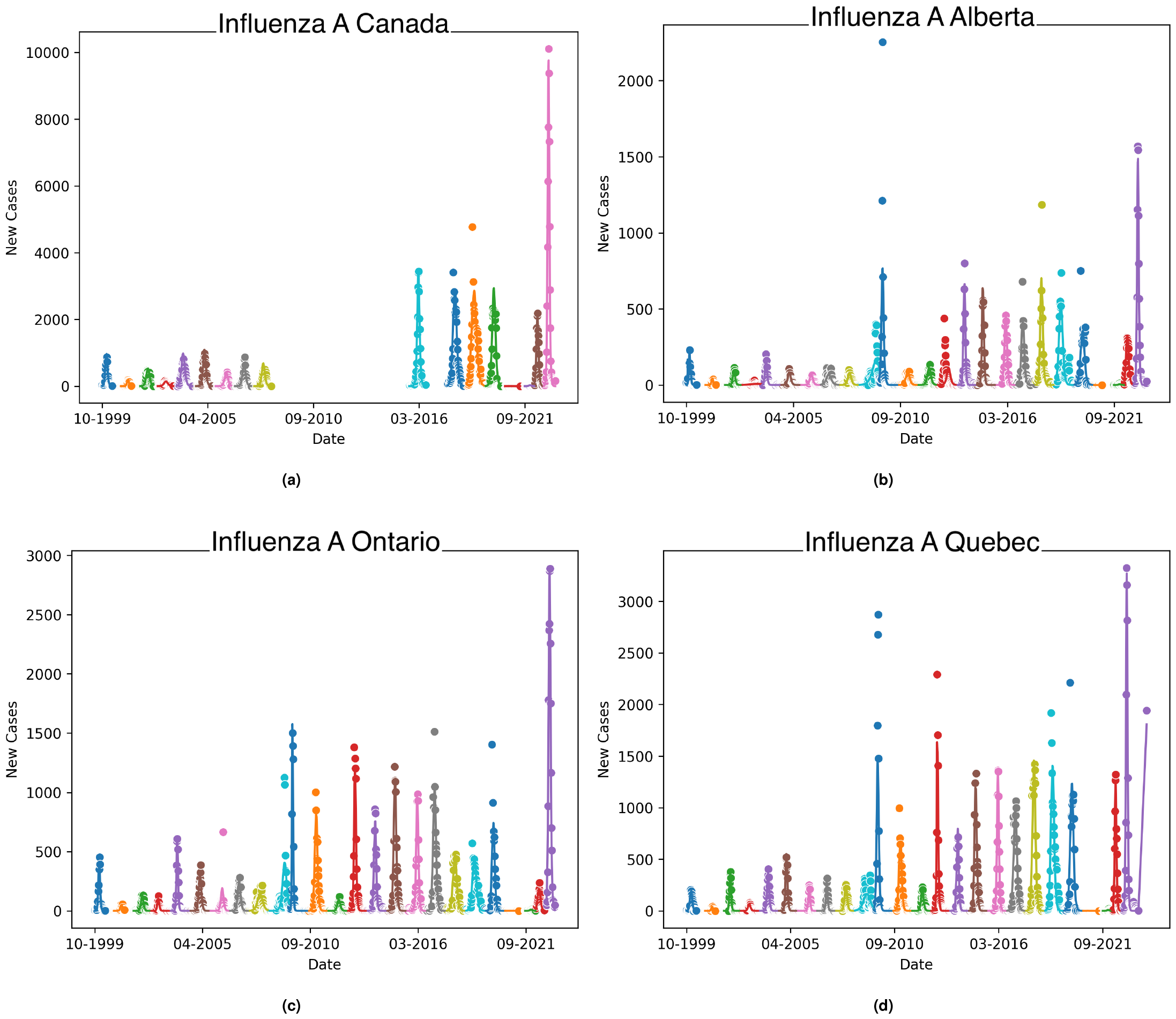
Influenza A data for Canada, Alberta, Ontario, and Quebec plotted per season starting from 1990 through 2023. The data is fit individually per season, as shown by the colour variation. Gaps show missing seasons from the dataset, or non-convergent fits.

Table 2 shows the estimates for *β, S*_0_, *R*_0_ and the Attack Rate for each flu season for Canada, Ontario, Alberta and Quebec for which at least one fitting method converged. We note that Canada’s FluWatch did not report provincial-level data for the 2020-2021 season, or for proceeding seasons. For these seasons, provincial level data was used when available (24–26), As such, this season is absent from analysis. Other missing rows of data are indicative of non-convergent fitting or large (*>* 2%) RRMSE. Of particular interest are seasons 2020 − 2021 and 2021 − 2022, during which NPIs for COVID-19 were highly used, including increased masking by the general public, widespread social distancing, and intermittent lockdowns. We see that the effect is largely seen in the base parameters through effective susceptible population size, *S*_0_. Which translates to decreases in *R*_0_ and Attack Rate.

**Table 2.** Fitted *β, S*_0_, *R*_0_ and computed Attack Rate for historical influenza A data for Canada, Ontario, Alberta and Quebec. Can see from this data that the effects of NPIs put in place for COVID-19 have quantitative and qualitative effects on the Influenza A. We see that on average we see approximately 5% decrease in the basic reproduction number of Influenza A, and we see that this mostly comes from a decrease in the effective susceptible population *S*_0_, rather than the contact rate, *β*. Note that Ontario experienced two waves of influenza A, one over 2021-2022 Winter, and one in Spring 2022.

| Season | Canada |  |  |  | Alberta |  |  |  |
| --- | --- | --- | --- | --- | --- | --- | --- | --- |
|  | beta | S0 | R0 | Attack Rate | beta | S0 | R0 | Attack Rate |
| 99-00 | 0.00016622 | 20462.8498 | 1.21473396 | 0.33483552 | 0.00151036 | 2457.57559 | 1.3256531 | 0.44755481 |
| 00-01 | 0.00084378 | 4028.24184 | 1.2139103 | 0.33065638 | 0.00541328 | 652.489663 | 1.26146686 | 0.3837101 |
| 01-02 | 0.00011566 | 27349.6485 | 1.12971228 | 0.22064108 | 0.00135953 | 2456.36191 | 1.19267561 | 0.30468599 |
| 02-03 | 0.00030219 | 10261.5702 | 1.10748155 | 0.18811571 | 0.0004846 | 5948.61466 | 1.02952665 | 0.05823518 |
| 03-04 | 6.05E-05 | 52142.6253 | 1.12735838 | 0.21760458 | 0.00100328 | 3427.25511 | 1.22803561 | 0.34724654 |
| 04-05 | 7.52E-05 | 42887.2548 | 1.15171921 | 0.25141863 | 0.0001697 | 17004.3483 | 1.03056289 | 0.06476137 |
| 05-06 | 0.00014047 | 22397.7092 | 1.12365913 | 0.21172432 | 0.00101366 | 3147.40347 | 1.1394265 | 0.23432425 |
| 06-07 | 0.00010339 | 30877.4116 | 1.140166 | 0.23537004 | 0.00037969 | 7983.6144 | 1.08259861 | 0.14970736 |
| 07-08 | 0.00010339 | 30877.4116 | 1.140166 | 0.23537004 | 0.00072213 | 4368.71252 | 1.12670388 | 0.21618121 |
| 08-09 | - | - | - | - | 0.00072213 | 4368.71252 | 1.12670388 | 0.21618121 |
| 09-10 | - | - | - | - | 0.00043458 | 9780.91446 | 1.5180509 | 0.5943478 |
| 10-11 | - | - | - | - | 0.00058206 | 5376.39581 | 1.11764071 | 0.20299028 |
| 11-12 | - | - | - | - | 0.00047632 | 6596.29444 | 1.12213486 | 0.20949767 |
| 12-13 | - | - | - | - | 0.00041514 | 7979.79024 | 1.18311704 | 0.29272945 |
| 13-14 | - | - | - | - | 0.00037289 | 9565.34304 | 1.2738693 | 0.39670902 |
| 14-15 | - | - | - | - | 0.00037808 | 9393.82048 | 1.26842327 | 0.39104772 |
| 15-16 | 3.41E-05 | 97157.9315 | 1.18415282 | 0.294231 | 0.00023391 | 14072.5729 | 1.17559655 | 0.28306968 |
| 16-17 | - | - | - | - | 0.00023164 | 14074.295 | 1.16433378 | 0.26844891 |
| 17-18 | 1.82E-05 | 172721.58 | 1.11996118 | 0.20692799 | 0.00018046 | 18464.4047 | 1.19006223 | 0.30154145 |
| 18-19 | 1.09E-05 | 280261.419 | 1.09060028 | 0.16248652 | 0.00010419 | 30140.5105 | 1.12155327 | 0.2100721 |
| 19-20 | 2.10E-05 | 150376.379 | 1.13046545 | 0.22182137 | 0.00014545 | 21557.9891 | 1.11987677 | 0.20638974 |
| 20-21 | 0.01075928 | 275.094253 | 1.05707739 | 0.11709196 | - | - | - | - |
| 21-22 | 0.0035854 | 916.891558 | 1.17408102 | 0.284589 | 0.02212772 | 148.882271 | 1.17658036 | 0.28451531 |
| 22* | 6.93E-05 | 48541.2115 | 1.2010877 | 0.3249449 | 0.00028839 | 11209.0558 | 1.1544847 | 0.26096192 |
| 22-23 | 2.09E-05 | 166948.38 | 1.24519128 | 0.36630147 | 0.00015764 | 22485.5027 | 1.26594304 | 0.38850824 |
| Season | Ontario |  |  |  | Quebec |  |  |  |
|  | beta | S0 | R0 | Attack Rate | beta | S0 | R0 | Attack Rate |
| 99-00 | 0.00074962 | 4851.85059 | 1.29894834 | 0.4230064 | 0.00077977 | 4342.60522 | 1.20937034 | 0.32530217 |
| 00-01 | 0.00296546 | 1138.75531 | 1.20604749 | 0.32103014 | 0.00541212 | 649.274594 | 1.25498398 | 0.37677228 |
| 01-02 | 0.00088892 | 3725.36893 | 1.18270203 | 0.29216692 | 0.00064844 | 5494.58375 | 1.27246434 | 0.39525457 |
| 02-03 | 0.00177442 | 1942.89855 | 1.23125333 | 0.35153767 | 0.00153609 | 2133.80177 | 1.17061285 | 0.27659946 |
| 03-04 | 0.00046549 | 7789.40159 | 1.29495522 | 0.41827243 | 0.00039401 | 8571.04925 | 1.20610585 | 0.32109775 |
| 04-05 | 0.0003318 | 9905.84902 | 1.1738567 | 0.28081599 | 0.00031742 | 10819.6541 | 1.22656879 | 0.34515459 |
| 05-06 | 0.00054947 | 5975.2032 | 1.17256237 | 0.27913414 | 0.00089931 | 3894.79047 | 1.25093969 | 0.37239322 |
| 06-07 | 0.00035166 | 9220.84317 | 1.15808148 | 0.25998643 | 0.00081906 | 4307.13253 | 1.25992366 | 0.38206704 |
| 07-08 | 0.00018104 | 16750.107 | 1.08298577 | 0.14954702 | 0.00040096 | 8148.89306 | 1.16690804 | 0.27173346 |
| 08-09 | 9.53E-05 | 32397.7244 | 1.10266733 | 0.18025097 | 7.70E-05 | 38451.7123 | 1.05740624 | 0.10710804 |
| 09-10 | - | - | - | - | - | - | - | - |
| 10-11 | 0.00014013 | 23577.9282 | 1.18001947 | 0.28877445 | 0.00015133 | 21651.0311 | 1.17014936 | 0.27602551 |
| 11-12 | 0.00086374 | 3730.03208 | 1.15063427 | 0.24987398 | 0.00050017 | 6561.96133 | 1.17216564 | 0.27861838 |
| 12-13 | 0.00014499 | 23590.2503 | 1.22153541 | 0.33955597 | 0.00017254 | 20963.9145 | 1.29181146 | 0.41490045 |
| 13-14 | 0.00022792 | 15057.8952 | 1.22570189 | 0.34435907 | 0.00021373 | 16006.0153 | 1.22177968 | 0.33962464 |
| 14-15 | 0.00011358 | 29328.1298 | 1.18968194 | 0.30103562 | 0.00011507 | 29182.9071 | 1.19935499 | 0.31292873 |
| 15-16 | 0.00015381 | 21998.8842 | 1.2084727 | 0.32393718 | 0.0001359 | 25391.9529 | 1.23238512 | 0.35179066 |
| 16-17 | 9.34E-05 | 34930.4781 | 1.16542693 | 0.26989898 | 6.85E-05 | 46505.5978 | 1.13827091 | 0.23275074 |
| 17-18 | 0.00011989 | 26279.2055 | 1.12525571 | 0.21463281 | 5.66E-05 | 57014.6818 | 1.15221044 | 0.25206623 |
| 18-19 | 0.0001122 | 27927.841 | 1.11908254 | 0.2056516 | 5.07E-05 | 62989.0653 | 1.14088207 | 0.23646914 |
| 19-20 | 0.0001419 | 23132.3505 | 1.17228102 | 0.27940252 | 5.73E-05 | 55742.2386 | 1.14034734 | 0.23568172 |
| 20-21 | 0.0093146 | 308.51246 | 1.02631054 | 0.06245293 | - | - | - | - |
| 21-22 | 0.01059945 | 311.293043 | 1.17840484 | 0.29000035 | 0.01365794 | 242.424461 | 1.18250705 | 0.29204132 |
| 22* | 0.00044435 | 7215.20374 | 1.14502913 | 0.26338778 | 0.00018216 | 19384.4053 | 1.26108525 | 0.3836769 |
| 22-23 | 5.23E-05 | 64596.1557 | 1.20684163 | 0.32451261 | 7.90E-05 | 45305.5685 | 1.27857975 | 0.40156215 |

We mark Spring/Summer 2022 as its own season as the changes in NPIs created conditions for a secondary flu season in 2022. This can be seen in Figures 3, and 4. We across all four jurisdictions a delayed Influenza A season in Spring of 2022.

**Fig. 3.**
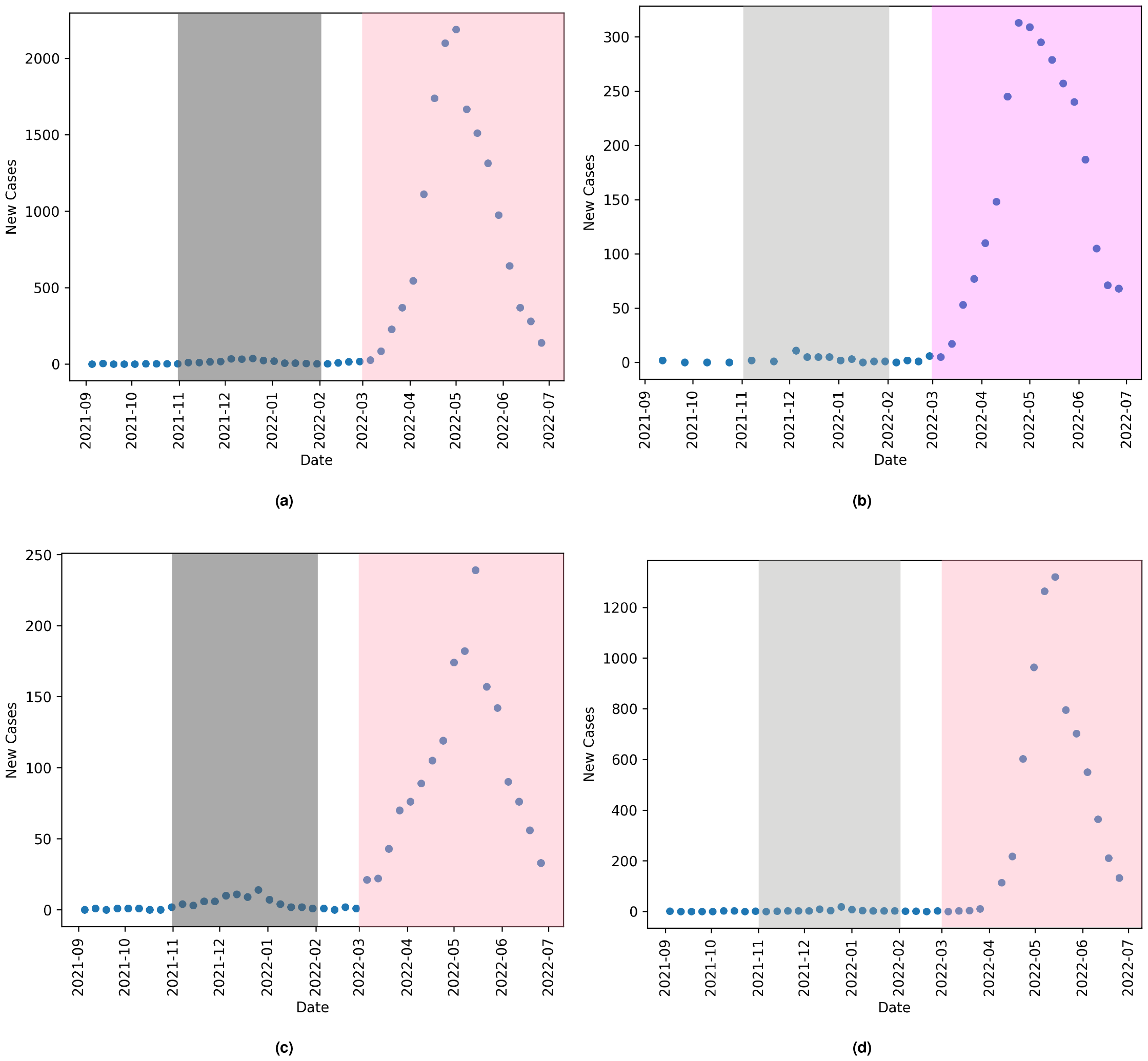
Influenza A reported cases for (a) Canada, (b) Alberta, (c) Ontario, and (d) Quebec. The grey shaded area shows the typical Influenza A season, and the pink shaded area shows the shifted outbreak in 2022. This was common across all three provinces and nationally.

**Fig. 4.**
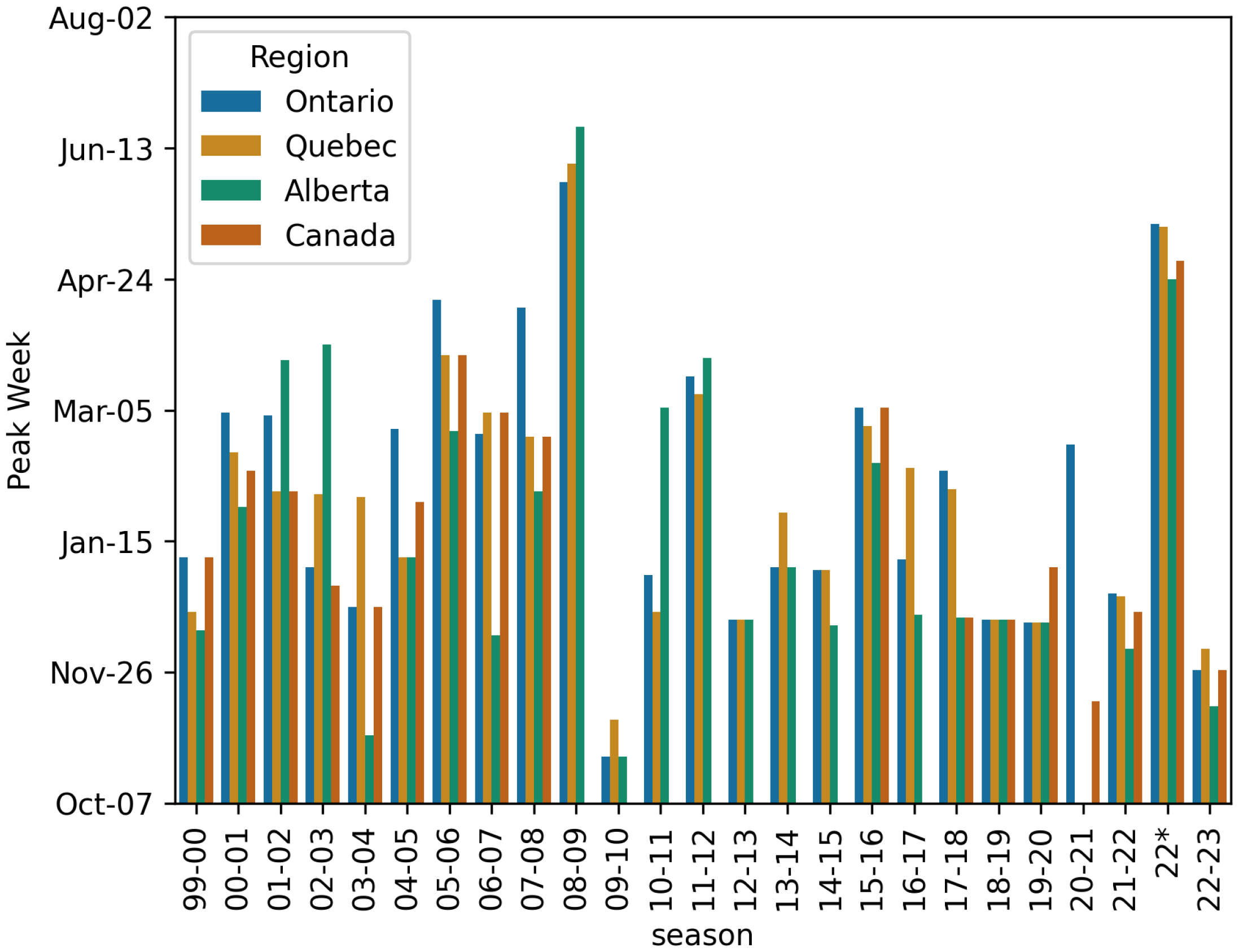
Plot showing the week with the most new Influenza A reports per season for each of the four studied jurisdictions. We see that in 2022 there was an abnormal outbreak of Influenza A in the spring. The year 08-09 is likely affected by the H1N1 pandemic which was officially declared in August 2009, straddling two influenza seasons.

Figure 5 shows the mean *R*_0_ and 95% confidence interval for this mean for season 99-00 through 18-19, deemed pre-COVID, and how the seasons 19-20, 20-21, 21-22, 22, and 22-23 compare. Similarly, Figure 6 shows the mean Attack Rate and a 95% confidence interval for this mean for the seasons 99-00 through 18-19. Also on the figure are point estimates of Attack Rate for the seasons 19-20 through 22-23. All mean values exclude the 09-10 season due to the effects of the H1N1 pandemic (28).

**Fig. 5.**
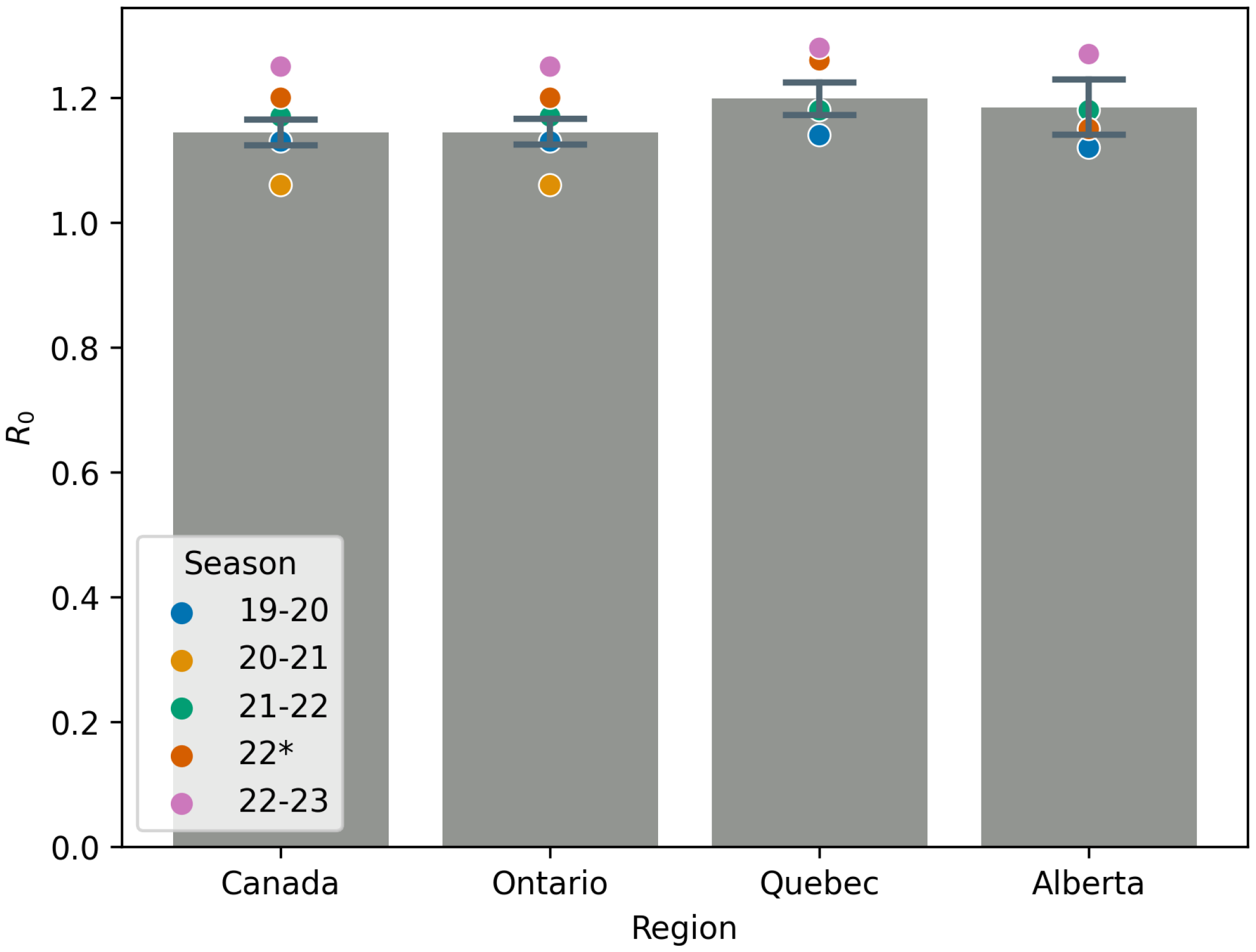
Mean *R*_0_ values for all seasons pre-COVID (i.e. seasons 99-00 through 18-19), the season 09-10 is excluded from any computation due to the H1N1 pandemic. Point estimates are given for the 19-20, 20-21, 21-22, 22, and 22-23 seasons for comparison.

**Fig. 6.**
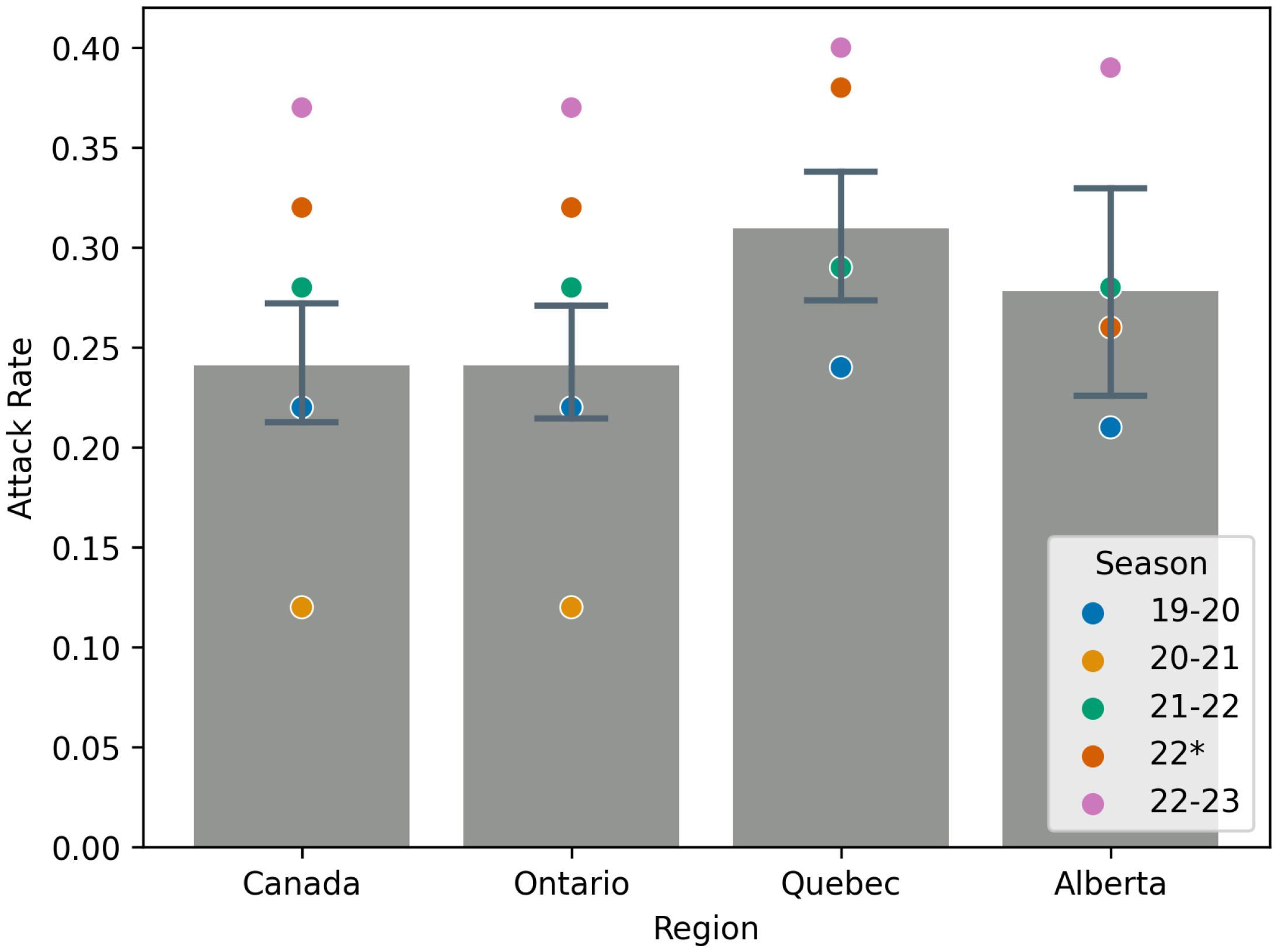
Mean Attack Rate values for all seasons pre-COVID (i.e. seasons 99-00 through 18-19), the season 09-10 is excluded from any computation due to the H1N1 pandemic. Point estimates are given for the 19-20, 20-21, 21-22, 22, and 22-23 seasons for comparison.

Table 3 gives mean values for *R*_0_, Attack Rate, and *S*_0_ for Influenza A across all seasons studied, with the exception of 2009 due to the H1N1 pandemic (27). This table largely quantifies information in Figures 5 and 6 and shows the relative change in value for seasons in which NPIs were in effect. We include the 22-23 season to highlight the substantial rebound in Influenza A ‘post’-COVID, when NPIs have been largely abandoned. Instead of a return to the mean, we see a more severe Influenza A season.

**Table 3.** Mean values of *R*_0_ and Attack Rate of Influenza A across all available pre-COVID seasons (with exception of the 2009-2010 season due to the H1N1 pandemic(27)) for Canada, Ontario, Alberta and Quebec. We also report the percent change from the mean for the 20-21, 21-22, and 22-23 seasons.

|  | R0 |  |  |  | Attack Rate |  |  |  |
| --- | --- | --- | --- | --- | --- | --- | --- | --- |
|  | Canada | Alberta | Ontario | Quebec | Canada | Alberta | Ontario | Quebec |
| Mean | 1.15±0.04 | 1.18±0.11 | 1.18±0.06 | 1.20±0.06 | 0.24±0.05 | 0.28±0.13 | 0.29±0.07 | 0.31±0.07 |
| % Change 20-21 | -7.70 | - | -13.29 | - | -51.37 | - | -78.40 | - |
| % Change 21-22 | 2.51 | -0.62 | -0.45 | -1.43 | 18.19 | 2.10 | 0.30 | -5.45 |
| % Change 22* | 4.87 | -2.49 | -3.27 | 5.12 | 34.95 | -6.35 | -8.90 | 24.22 |
| % Change 22-23 | 8.72 | 6.93 | 1.96 | 6.57 | 52.13 | 39.42 | 12.24 | 30.01 |

The pre-COVID mean values of *R*_0_ and the attack rate are similar to those found in the literature (19, 29–31).

## 5. Discussion

The results show that the effective initial susceptible population size is greatly affected by NPIs. While *S*_0_ and *β* are correlated in the expression for *R*_0_, the additional information at hand, like the number of tests conducted and the cumulative number of infections in one season, give us bounds on *S*_0_ so that a model parameterized with the same *R*_0_, but different *S*_0_ will not yield the same fit.

With less biased and more randomized testing of influenza case data, this information could readily be applied to an entire population to monitor the effectiveness of NPIs during an outbreak of a novel infectious disease with similar transmission routes to the flu. As it stands, the raw parameters are subject to limitations of available data.

Due to these limitations, we focus on the relative change in epidemiological parameters of seasonal influenza A in each jurisdiction of study. In the 2020-2021 flu season, we see large changes (50% decrease) in the attack rate for influenza and about a 7% decrease in the reproduction number for influenza nation-wide. Further suggesting that when forecasting and modeling disease transmission during periods of wide-spread societal changes, the effective population number is incredibly important and cannot be assumed to be the entire population. Using a known disease with similar routes of transmission may help guide estimates for effective population size during future outbreaks of novel infectious diseases.

We note that in Table 3, we see that by Spring 2022, the behavioural changes induced by the COVID-19 pandemic largely fell out of favour nationally leading to a moderate rebound of influenza A. We see specifically in Ontario, where the data exists, that the reduction in *R*_0_ and the Attack Rate are substantially lower than in the 20-21 season.

These values also give measures of adherence to NPIs. We see, by comapring the % Change in the 21-22, 22, and 22-23 seasons that more Ontarians likely continued to practice person NPIs like masking and distancing. Another factor that can impact the milder rebound of influenza could be vaccine up-take of the flu vaccine in different jurisdictions.

Because they are inversely correlated, we see in our fitting that when *S*_0_ sees a significant decrease, *β*, the contact rate, shows a significant increase. The fact that *R*_0_ was lower during the 2020-2021 season shows that the increase in *β* is not exactly proportional to the decrease in *S*_0_. Our interpretation is that there are effectively fewer individuals in the population but they are generally those with higher contacts. This may inform estimates of effective contact rates of the subsets of the population who are considered ‘essential workers’ during a pandemic.

The data shown in Figure 2 shows, particularly in the 2021-2022 season, that temporary regional lockdowns had minimal effect on influenza. This could be due to the timing of the lockdowns compared to the flu season of that year, or the relative length of flu season against the length and scope of lockdowns. This suggests that it was the social behaviours like masking and long-term policy and behavioural changes that drove a reduction in influenza A cases.

Future work includes expanding this framework and pipeline to other endemic, airborne infections in Canada to see if different pathogens are affected differently by NPIs. This work could help inform how certain policies will affect pathogens with differing properties. This could lead to targeted, effective NPIs being put in place once certain characteristics of infectious diseases are known.

We would also like to expand to different jurisdictions, perhaps all provinces and territories of Canada and other countries. Here we focus on three large provincial jurisdictions to highlight that this fitting pipeline can be extended to the provincial level and produces consistent results.

Another curiosity is the increase in the reproduction number and the attack rate in the 2022-2023 season when NPIs and associated policies were phased out by the population at large. This may be tied to a possible decrease in uptake of the flu shot(32), a reduction in cross-immunity through several seasons without exposure to Influenza A, or cross-infection with COVID-19 (33). While there are many open questions about the mechanics of this rebound that we aim to study in the near future, we hope that these estimates of the effects on NPIs on an endemic infection like influenza A can help modeling and forecasting in the future.

## Data Availability

The data are all publicly available through Canada, Ontario, Quebec and Alberta health portals. All portals are cited in the text.

## References

1. SC Anderson, et al., Quantifying the impact of covid-19 control measures using a bayesian model of physical distancing. PLoS computational biology 16, e1008274 (2020).

2. A Aleta, et al., Modelling the impact of testing, contact tracing and household quarantine on second waves of covid-19. Nat. Hum. Behav. 4, 964–971 (2020).

3. A Karaivanov., SE Lu, H Shigeoka, C Chen, S Pamplona, Face masks, public policies and slowing the spread of covid-19: Evidence from canada. J. Heal. Econ. 78, 102475 (2021).

4. A Peng, S Bosco, A Tuite, A Simmons, D Fisman, Impact of community masking on sars-cov-2 transmission in ontario after adjustment for differential testing by age and sex. medRxiv pp. 2023–07 (2023).

5. IR Moyles, JM Heffernan, JD Kong, Cost and social distancing dynamics in a mathematical model of covid-19 with application to ontario, canada. Royal Soc. open science 8, 201770 (2021).

6. MI Betti, JM Heffernan, A simple model for fitting mild, severe, and known cases during an epidemic with an application to the current sars-cov-2 pandemic. Infect. Dis. Model. 6, 313–323 (2021).

7. PHAo Canada, Government of canada (2021).

8. M Koutsakos, AK Wheatley, K Laurie, SJ Kent, S Rockman, Influenza lineage extinction during the covid-19 pandemic? Nat. Rev. Microbiol. 19, 741–742 (2021).

9. R Ross, Some quantitative studies in epidemiology. Nature 87, 466–467 (1911).

10. G Chowell, L Sattenspiel, S Bansal, C Viboud, Mathematical models to characterize early epidemic growth: A review. Phys. life reviews 18, 66–97 (2016).

11. R Casagrandi, L Bolzoni, SA Levin, V Andreasen, The sirc model and influenza a. Math. biosciences 200, 152–169 (2006).

12. G Nakamura, B Grammaticos, M Badoual, Vaccination strategies for a seasonal epidemic: a simple sir model. Open Commun. Nonlinear Math. Phys. 1 (2021).

13. B Sara, Z Omar, T Abdessamad, R Mostafa, F Hanane, Parameters’ estimation, sensitivity analysis and model uncertainty for an influenza a mathematical model: case of morocco. Commun. Math. Biol. Neurosci. 2020, Article–ID (2020).

14. C Reed, et al., Estimating influenza disease burden from population-based surveillance data in the united states. PloS one 10, e0118369 (2015).

15. SL Wu, et al., Substantial underestimation of sars-cov-2 infection in the united states. Nat. communications 11, 4507 (2020).

16. L Kalachev, EL Landguth, J Graham, Revisiting classical sir modelling in light of the covid-19 pandemic. Infect. Dis. Model. 8, 72–83 (2023).

17. D Osthus, KS Hickmann, PC Caragea, D Higdon, SY Del Valle, Forecasting seasonal influenza with a state-space sir model. The annals applied statistics 11, 202 (2017).

18. S Boonpatcharanon, JM Heffernan, H Jankowski, Estimating the basic reproduction number at the beginning of an outbreak. Plos one 17, e0269306 (2022).

19. AR Tuite, DN Fisman, JC Kwong, AL Greer, Optimal pandemic influenza vaccine allocation strategies for the canadian population. PloS one 5, e10520 (2010).

20. P Van den Driessche, J Watmough, Reproduction numbers and sub-threshold endemic equilibria for compartmental models of disease transmission. Math. biosciences 180, 29–48 (2002).

21. PHAo Canada, Government of canada (2024).

22. ON, Bjørnstad., C Viboud, Timing and periodicity of influenza epidemics. Proc. Natl. Acad. Sci. 113, 12899–12901 (2016).

23. S Edlund, et al., Comparing three basic models for seasonal influenza. Epidemics 3, 135–142 (2011).

24. O Public Health, Ontario respiratory virus tool (2024).

25. dQ Institut national de santé publique, Archives influenza: Inspq (2024).

26. A Government of, Respiratory virus dashboard (2024).

27. SM Moghadas, NJ Pizzi, J Wu, SE Tamblyn, DN Fisman, Canada in the face of the 2009 h1n1 pandemic. Influ. Other Respir. Viruses 5, 83–88 (2011).

28. M Patel, A Dennis, C Flutter, Z Khan, Pandemic (h1n1) 2009 influenza. Br. journal anaesthesia 104, 128–142 (2010).

29. D He, J Dushoff, R Eftimie, DJ Earn, Patterns of spread of influenza a in canada. Proc. Royal Soc. B: Biol. Sci. 280, 20131174 (2013).

30. PR Lagace-Wiens, E Rubinstein, A Gumel, Influenza epidemiology—past, present, and future. Critical care medicine 38, e1–e9 (2010).

31. C Sikora, et al., Transmission of pandemic influenza a (h1n1) 2009 within households: Edmonton, canada. J. clinical virology 49, 90–93 (2010).

32. H Lymon, Declines in influenza vaccination coverage among health care personnel in acute care hospitals during the covid-19 pandemic—united states, 2017–2023. MMWR. Morb. Mortal. Wkly. Rep. 3 72 (2023).

33. SS Lee, C Viboud, E Petersen, Understanding the rebound of influenza in the post covid-19 pandemic period holds important clues for epidemiology and control. Int. J. Infect. Dis. 122, 1002–1004 5 (2022).

